# Intention to breastfeed and association with subsequent breastfeeding duration, a linked population level routine data study - The Born in Wales cohort 2018-2021

**DOI:** 10.1101/2022.12.13.22283407

**Authors:** H Jones, M Seaborne, M Mhereeg, M James, N Kennedy, A Bandyopadhyay, S Brophy

## Abstract

**Objective:** The World Health Organisation recommends exclusive breastfeeding for the first six months of life. This study aimed to examine 1) If intention to breastfeed is associated with higher levels of breastfeeding, and 2) If the pandemic impacted the intention to breastfeed and the breastfeeding rates in Wales.

**Methods:** This was a cohort study utilising routinely collected, linked healthcare data from the Secure Anonymised Information Linkage databank (SAIL). SAIL combines data from the National Community Child Health (NCCH) and Maternal Indicators (MIDS) datasets. All women that gave birth between 2018 and 2021 recorded in the NCCH dataset and MIDS dataset were included.

**Results:** Intention to breastfeed was associated with being 27.5 times more likely to continue to breastfeed for six months compared to those who did not intend to breastfeed (OR 27.5, 95% CI: 24.8-30.5). 57.8% of expectant mothers intended to breastfeed pre-pandemic and 58.7% intended to breastfeed in 2020. Exclusive breastfeeding rates to six months were 16.6% pre-pandemic, 20.5% in 2020 and 19.7% in 2021. Those aged 30-39 were 3.91 times more likely to breastfeed.

**Conclusion:** The biggest predictor of exclusive breastfeeding at six months was intention to breastfeed. During the pandemic those who intended to breastfeed were more likely to breastfeed exclusively for six months compared to before and after the pandemic, suggesting time at home may facilitate breastfeeding. Therefore, targeted interventions during pregnancy planning and early pregnancy to promote and encourage intention to start breastfeeding and promoting home working could improve breastfeeding rates and duration.

## Introduction

The considerable health and social benefits of breastfeeding are already well-established [1]. Breastfeeding provides both short-term and long-term physical, economic and environmental improvements for young children, women, and society. Benefits to breastfed babies include fewer infections, increased intelligence, and protection against obesity and diabetes. Exclusively breastfed babies have been demonstrated to have a higher intelligence quotient (IQ) and may have higher mathematical abilities than artificially fed babies (those fed with bottle milk, formula milk or alternative methods) [2]. They are also less prone to infections, asthma, other atopy, and allergic diseases. Moreover, breastfeeding is beneficial for mothers as research indicates that it reduces the risk of breast, uterine and ovarian cancer, reduces postpartum bleeding, and is beneficial in promoting postpartum weight loss [2]. Increasing the rates of breastfeeding on a worldwide scale could help prevent 823,000 deaths per year in children under five years old and prevent 20,000 maternal deaths per year from breast cancer [3]. To recognise the advantages of breastfeeding, political support and financial investment are essential to protect, promote, and support breastfeeding [3].

The current World Health Organisation (WHO) recommendations indicate that mothers should provide exclusive breastfeeding to their babies during the first six months of life [4]. However, the United Kingdom (UK) has one of the lowest breastfeeding rates in the world. In fact, 80% of babies are breastfed at birth but only 1% are exclusively breastfed for six months in the UK [5]. These breastfeeding rates are lower among women in areas of higher deprivation and health inequalities [6]. Breastfeeding rates in Wales have remained consistent despite investment in services and UNICEF baby friendly initiative accreditation [5]. It is acknowledged that this is a multi-faceted issue relating to population health, initiation, and maintenance of breastfeeding. It is important to note that breastfeeding rates are influenced by various socio-economic factors, religion, education, and support services available. [6]

Factors that influence women’s breastfeeding decisions can include breastfeeding intention, breastfeeding self-efficacy and social support [7]. Breastfeeding promotion strategies conducted by midwives often include social support, however, there is minimal research surrounding intention to breastfeed and self-efficacy and whether this impacts future breastfeeding behaviour.

In 2020, the COVID-19 pandemic and concerns about transmission of the disease contributed to higher rates of breastfeeding interruption [8]. Research has found that the risk of transmission from infected mothers to their offspring is extremely low [8]. Furthermore, available data [8] showed that the lack of support for lactating mothers during the pandemic contributed to breastfeeding cessation worldwide. Few strategies were proposed to overcome this issue.

In the year 2020, research found that a third of women who planned to breastfeed reported that they did not receive help with positioning the infant [9]. Additionally, one in four women perceived that they did not get enough support with feeding in the hospital [9]. This could have been due to the increased burden on healthcare systems and pressure placed on healthcare professionals to discharge mothers sooner to minimise infection risks, resulting in fewer opportunities to support mothers with infant feeding. It is unclear whether rates of breastfeeding changed again after the periods of lockdowns and restrictions in Wales and upon returning to normal day-to-day activity.

The protection, promotion, and support of breastfeeding is a priority for public health. Therefore, encouraging optimal breastfeeding practices including breastfeeding initiation within one hour after birth, exclusive breastfeeding for six months, and continuation of breastfeeding for at least two years once nutritious and safe complementary foods are introduced at six months is crucial for meeting the Sustainable Development Goals by 2030 [10]. For example, goals 2 and 3 are concerned with hunger, health, and well-being [10]. Breastfeeding is a vital source of nutrition that can save children’s lives and contribute to improved health outcomes for children and mothers.

The aims of the study are to 1) investigate the changes in breastfeeding rates from 2018 to 2021 in Wales using population-level data in the SAIL databank, and 2) examine if pregnant women intending to breastfeed do so. Understanding what factors have an impact on breastfeeding up to the recommended six months will help develop more targeted interventions to promote and support breastfeeding.

## Materials and Methods

### Study Design and Setting

This was a cohort study utilising routinely collected anonymised population-level linked data from the Secure Anonymised Information Linkage (SAIL) databank [11,12]. Data sources include the National Community Child Health (NCCH) and Maternal Indicators (MIDS) datasets. All women in Wales included in the NCCH Breastfeeding and MIDS data were deemed eligible for analysis; any stillbirths were excluded. In cases where women had multiple reported births, all births were selected as the mother’s outcome of breastfeeding and intention to feed at each birth is independent of the decisions at another birth. Breastfeeding rates were compared from 2018 to 2021.

### Data Sources and Linkage

Analysis was undertaken using anonymised population-scale, individual level linked routinely collected national-scale data available in the SAIL Databank [11,12]. This anonymously links a wide range of person-based data using a unique, encrypted personal identifier. The linkage in this study includes NCCH Births, NCCH Breastfeeding and MIDS data. The NCCH comprises information pertaining to birth registration, monitoring of child health examinations, and immunisations. The MIDS data contains data relating to pregnant women at their initial antenatal assessments, and mother and baby (or babies) for all births. These records were linked at the individual level for all women known to be pregnant in Wales between 2018 and 2021.

Data contained in the breastfeeding dataset included the outcome. The outcomes are categorised in SAIL into exclusively breastfeeding, combined - predominantly breastfeeding, or combined - partial breastfeeding and artificial milk. The collection times were categorised into first feed, ten days, six weeks and six months. The NCCH breastfeeding dataset was linked to the MIDS dataset to see whether a mother’s intention to breastfeed resulted in an outcome of exclusive milk feeding. They were asked whether they intended to breastfeed and only a yes or no response was noted. All missing (NA) outcome values were removed and regression undertaken with the MIDS intention to breastfeed yes or no binary. Intention to feed from MIDS is compared to responses to the Born in Wales survey [13] that also asked how expectant mothers were planning to feed their baby to see if findings were consistent.

### Ethical Approval

The data held in the SAIL databank in Wales are anonymised. All data contained in SAIL has the permission from the relevant Caldicott Guardian or Data Protection Officer. This study is part of Born in Wales which was approved by the Health Research Authority on 16/06/2021, protocol number RIO 030-20.

### Statistical Analysis

Descriptive statistics were conducted on rates of breastfeeding per year from 2018 to 2021. Binary logistic regression models were utilised to examine the impact of the explanatory variables age group, ethnicity, and intention to breastfeed on breastfeeding rates, reporting odds ratios (OR) with 95% confidence intervals and significance level accepted at p<0.05. For intention, ‘yes’/’no’ responses were coded as 1 and 0 respectively and that there was no other outcome to be coded. The reference groups were those aged <18, white ethnicity, and those intending to breastfeed. The data handling and preparation for the descriptive statistics, logistic regression and survival analysis were performed in RStudio [14]. Final data preparation specific to these analyses such as setting the reference groups was performed in RStudio. Descriptive statistics, logistic regression and survival analysis were performed in RStudio.

## Results

A total of 122,387 babies born to 106,022 mothers were identified from 2018 to 2021. Excluding those babies born outside of Wales (N=3,172), stillbirths (N=494) and those having no data available for breastfeeding outcome (N=1,721), the cohort for analysis of breastfeeding rates comprised N=117,000 babies. A further N=22,055 babies were excluded because of NA values for maternal age, intention to breastfeed and breastfeeding outcome meaning the regression analysis was conducted on N=94,945 babies (Figure 1 describes the participants in the cohort). Most of the women were aged between 30-39 (46.7%) and between 25-29 years (29.7%). The majority were White (81.9%) and the majority intended to breastfeed (58.1%), see Table 1. The study population had higher numbers of women who did not intend to breastfeed (40%) compared to the all Wales database (33%).

**Table 1.**
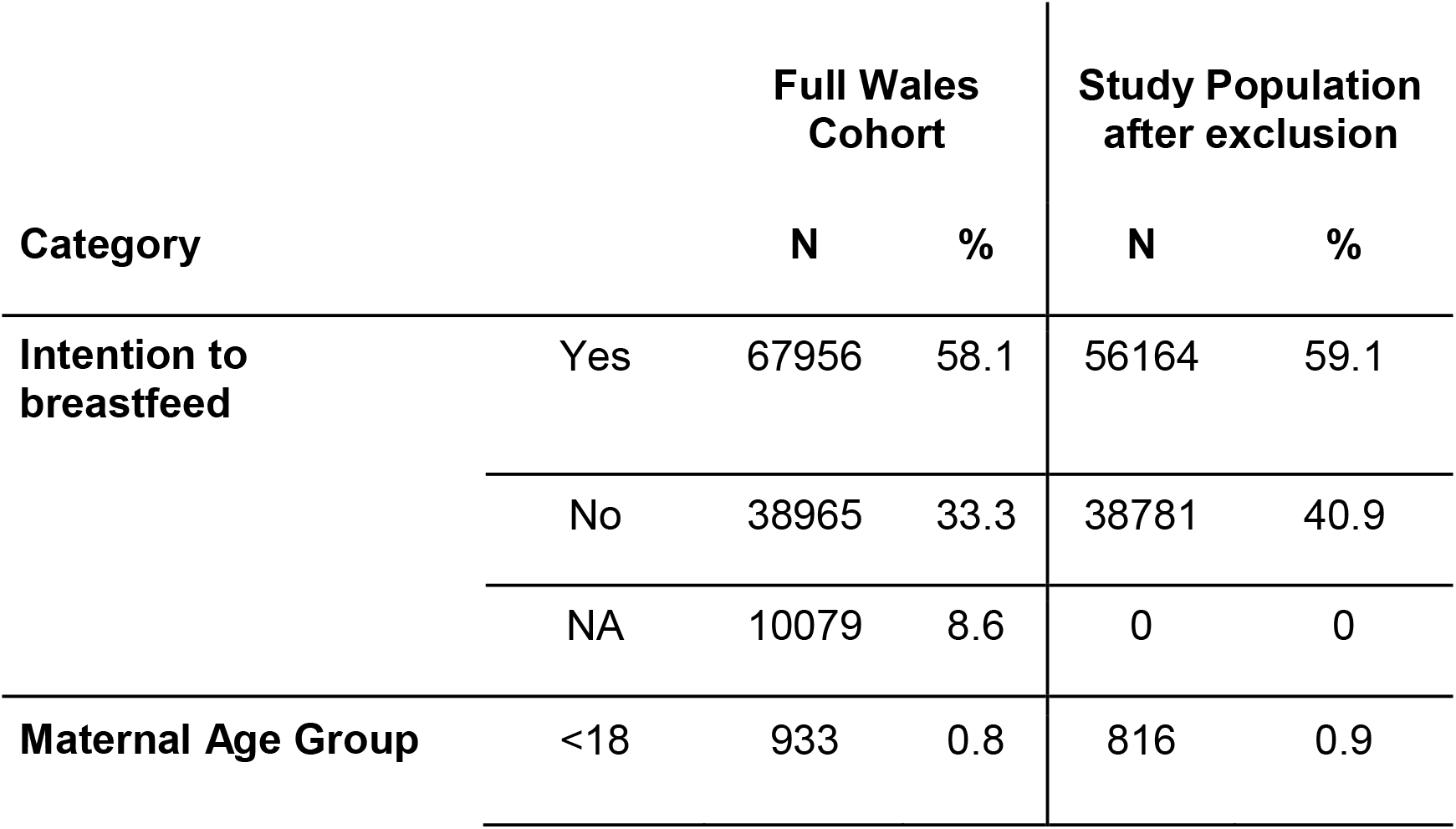
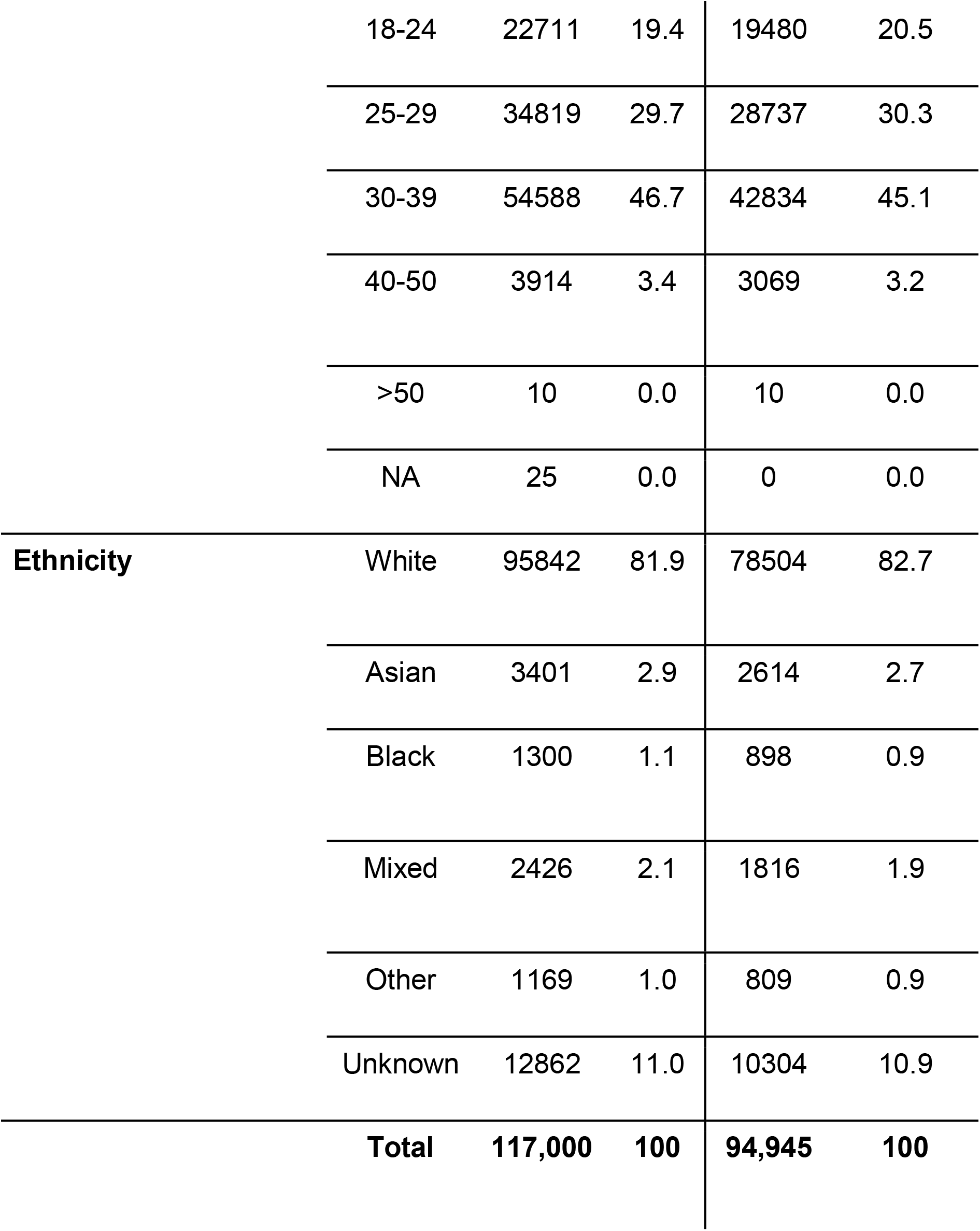
Demographics of the study cohort

| Category |  | Full Wales Cohort |  | Study Population after exclusion |  |
| --- | --- | --- | --- | --- | --- |
|  |  | N | % | N | % |
| Intention to breastfeed | Yes | 67956 | 58.1 | 56164 | 59.1 |
|  | No | 38965 | 33.3 | 38781 | 40.9 |
|  | NA | 10079 | 8.6 | 0 | 0 |
| Maternal Age Group | <18 | 933 | 0.8 | 816 | 0.9 |
|  | 18-24 | 22711 | 19.4 | 19480 | 20.5 |
|  | 25-29 | 34819 | 29.7 | 28737 | 30.3 |
|  | 30-39 | 54588 | 46.7 | 42834 | 45.1 |
|  | 40-50 | 3914 | 3.4 | 3069 | 3.2 |
|  | >50 | 10 | 0.0 | 10 | 0.0 |
|  | NA | 25 | 0.0 | 0 | 0.0 |
| **Ethnicity** | White | 95842 | 81.9 | 78504 | 82.7 |
|  | Asian | 3401 | 2.9 | 2614 | 2.7 |
|  | Black | 1300 | 1.1 | 898 | 0.9 |
|  | Mixed | 2426 | 2.1 | 1816 | 1.9 |
|  | Other | 1169 | 1.0 | 809 | 0.9 |
|  | Unknown | 12862 | 11.0 | 10304 | 10.9 |
|  | **Total** | **117,000** | **100** | **94,945** | **100** |

**Figure 1.**
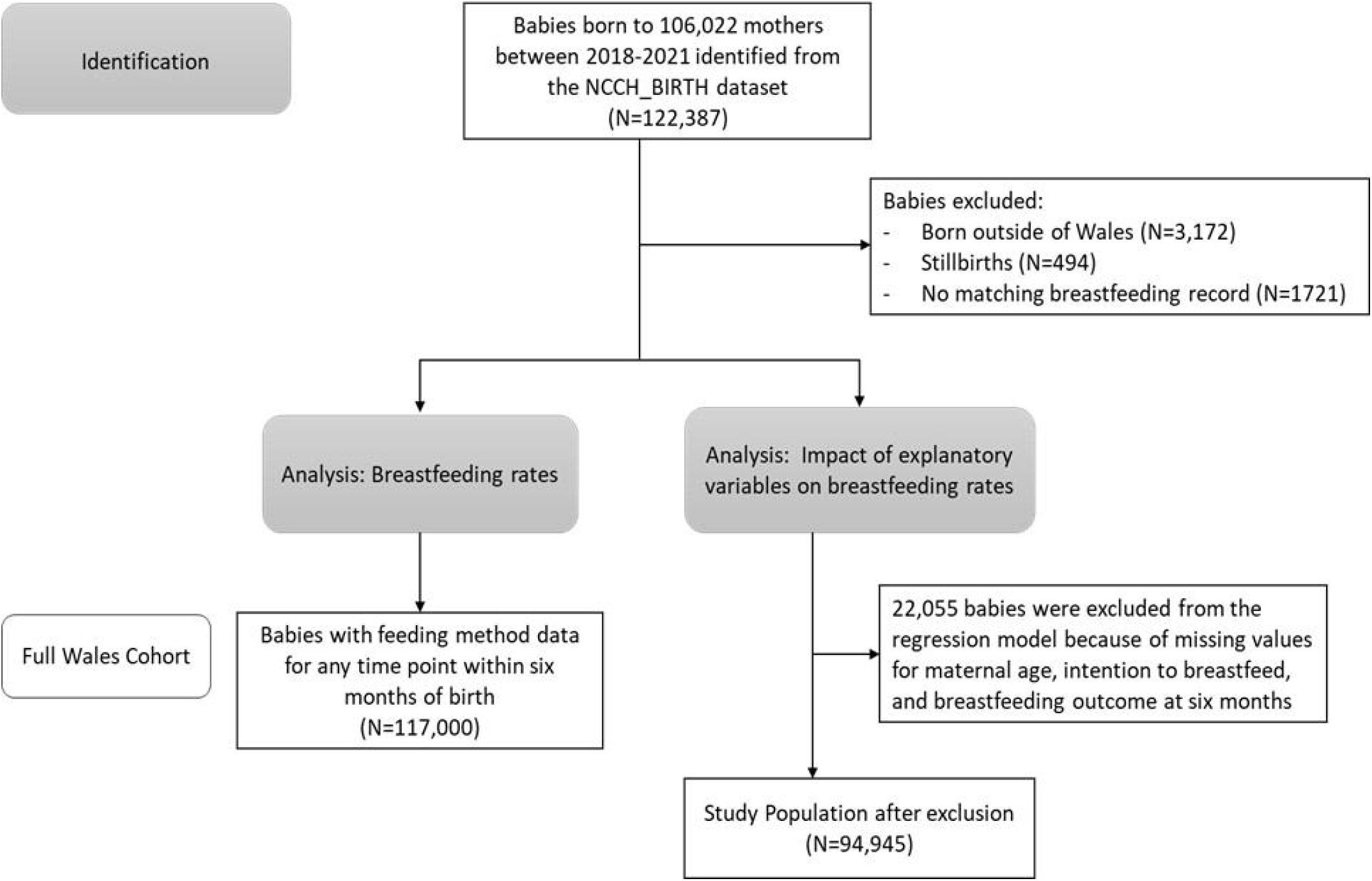
Study flowchart explaining how the cohort was identified

**Figure 2.**
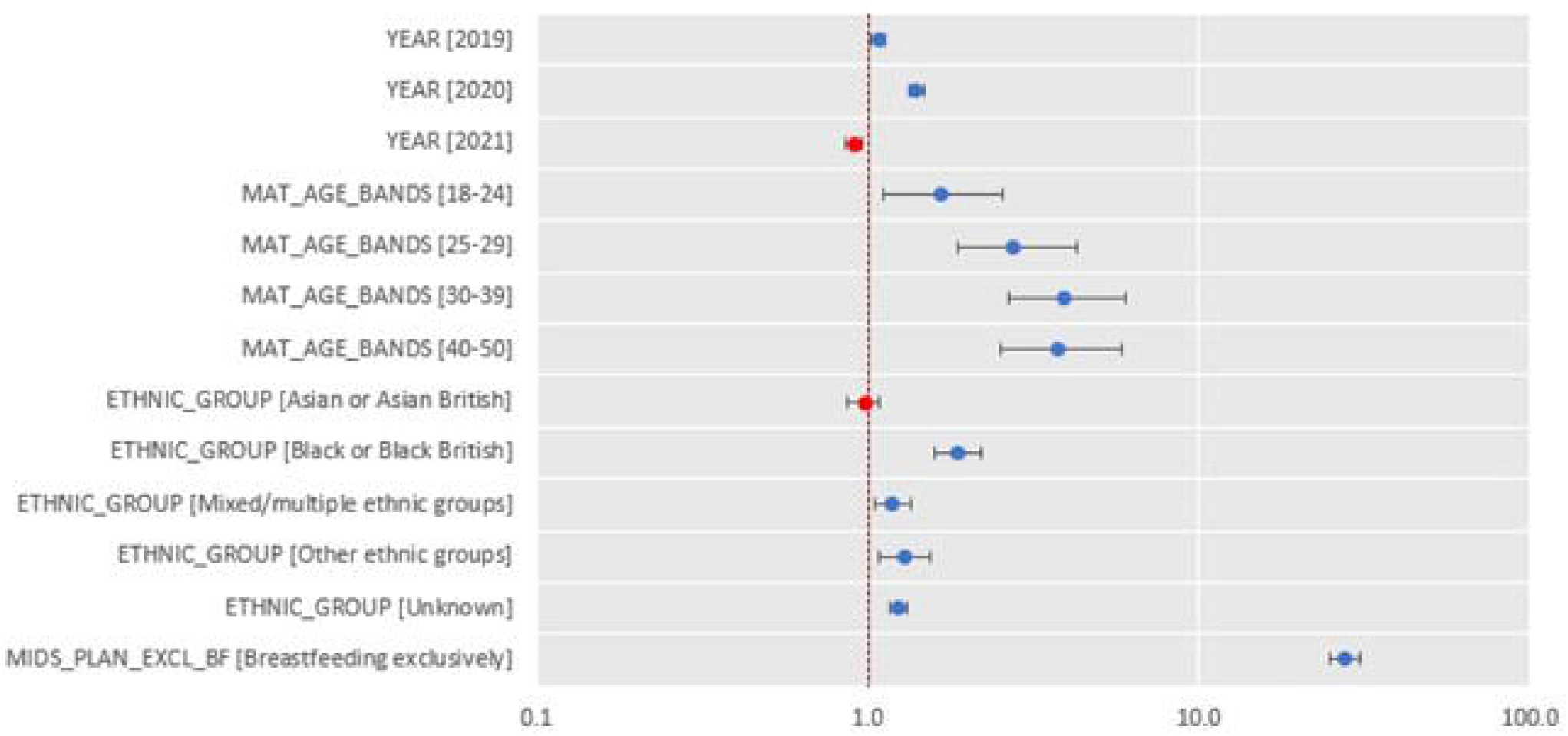
Logistic regression for exclusive breastfeeding for six months

### Rates of breastfeeding pre-pandemic, 2020 and 2021

Across the four years, between 59.9% to 61.9% of women breastfed their baby for their first feed. This decreased to between 35.1% to 37.3% breastfeeding at ten days, 25.4% to 28.5% at six weeks and those exclusively breastfeeding were lowest at six months at 16.6% to 20.6%. Between 11.7% to 14% at least partially breastfed at ten days, 7.5% to 8.4% at six weeks and 3.4% to 4.9% at six months. The other forms of feeding including artificial milk and a combination of breastmilk and artificial milk were mostly adopted after the first feed. The biggest difference was pre-pandemic and 2020 for breastfeeding to six months 16.6% compared to 20.6% (difference: 3.99% (CI 3.35-4.62%)).

### Examining the impact of intention to breastfeed, age, and ethnicity on breastfeeding outcome

The study found that the number of women intending to breastfeed did not differ during the pandemic (57.8% of expectant mothers intended to breastfeed before the pandemic compared to 58.7% in 2020 (difference: −0.88 (01.58,-0.18%))). Logistic regression was conducted to examine the variations in breastfeeding rates by intention to breastfeed, adjusted for age, year and ethnicity. The model explained 12% (R^2^ Tjur) of the variance in exclusive breastfeeding. Mothers in 2020 were 1.40 times more likely to breastfeed for six months compared to mothers pre-pandemic (OR=1.40, 95% CI 1.33-1.48, p < 0.001). Those who intended to breastfeed were 27.47 times more likely to breastfeed for six months compared to those who did not intend to breastfeed (OR=27.47, 95% CI 24.79-30.54, p <0.001). Those aged 30-39 were 3.91 times more likely to breastfeed for six months compared to those aged <18 (OR=3.91, 95% CI 2.65-6.04, p < 0.001), see Table 2 and Figure 3 below. Those of Black or Black British ethnic group were 1.88 times more likely to breastfeed for six months compared to White ethnic group (OR=1.88, CI 95% 1.60-2.20, p < 0.001).

**Table 2.**
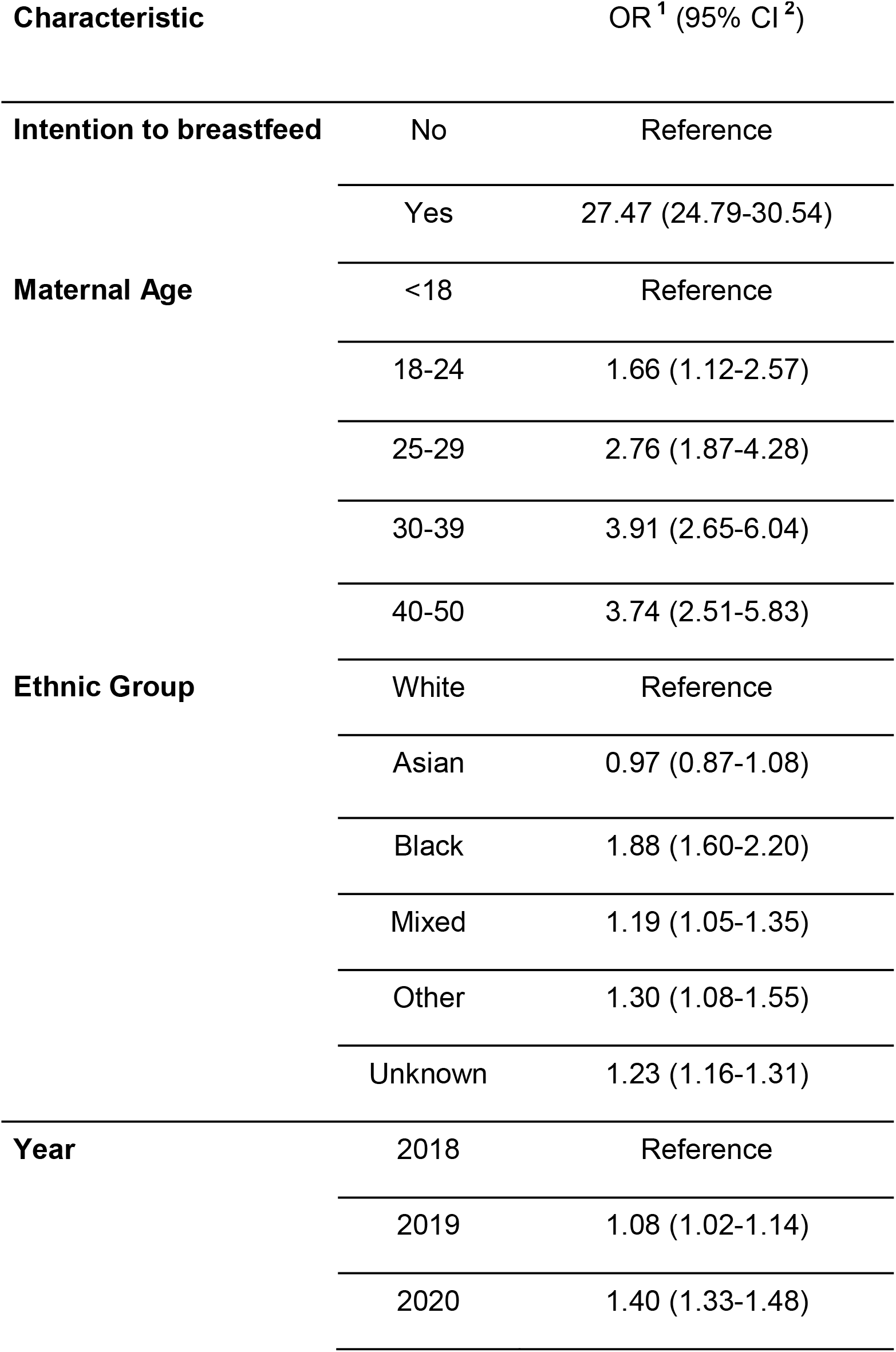
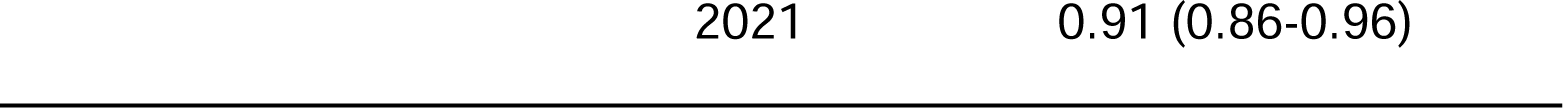
Binary Logistic Regression analysis of factors associated with breastfeeding outcome at six months. OR - Odds Ratio. CI - Confidence Interval.

### Intention to breastfeed in Born in Wales and MIDS

Of 258 women who completed a survey during pregnancy and had a record in MIDS, 208 (80%) reported an intention to breastfeed. Of these, 88% (183/208) reported the intention to breastfeed when asked again when being checked in for the birth (on the MIDS form). 12% (25/208) had changed their mind and now intended to not breastfeed or were no longer sure. If a woman reported the intention to not breastfeed on the survey during pregnancy, they changed their mind and reported they would like to breastfeed in less than 9% of cases. This suggests among a survey population, the initial intention to breastfeed/not breastfeed only changes for about 10% of women and so does not change easily.

## Discussion

The findings indicate that approximately 58% of expectant mothers intend to breastfeed and approximately 60% of mothers in Wales breastfeed for their first feed across the four years. However, only 16.6% to 20.6% continue to exclusively breastfeed up to the WHO recommended duration of six months. The rates of breastfeeding to six months were highest during 2020 when the COVID-19 pandemic was at its height although intention did not significantly change during 2020. This is in contrast to previous research that indicated COVID-19 negatively affected breastfeeding [9]. The rates of breastfeed to 6 months are now returning to pre pandemic levels.

A systematic review to identify factors influencing exclusive breastfeeding up to six months has previously been conducted [15]. Exclusive breastfeeding up to six months was reportedly affected by maternal working status, breastfeeding knowledge, delivery mode, perception of insufficient human milk, mothers’ infant feeding attitude, breastfeeding self-efficacy, and intention [15]. Breastfeeding up to six months is associated with working status and breastfeeding knowledge. Supporting women to visualise how they might continue to breastfeed given work commitments, e.g. where it is feasible to work from home, and improving knowledge early on in pregnancy could help to improve intention to breastfeed for longer.

The timing of asking about intention to breastfeed is important to examine. If intention was asked at the start of the pregnancy and this led to a higher uptake of exclusive breastfeeding for six months, it would point to the need to influence women’s perceptions of breastfeeding even prior to getting pregnant. For example, a social media campaign and advertising. It is rare to see adverts promoting breastfeeding outside of a healthcare setting but there are adverts for artificial milk. It would be interesting to have an in-depth study of women’s reasons for and against breastfeeding and reasons why they stop and link these to the data.

## Limitations

The study utilises health care data for pregnant women and new mothers in Wales including the maternity and child health data; thus providing a national perspective of breastfeeding rates established on a total population cohort, enabling the findings to be generalisable. However, it must be noted that it is difficult to fully know whether the data that was available on breastfeeding is reliable. The quality of data may be influenced by collection methods and how often health visitors are able to visit families to ask questions about methods of feeding and how long they breastfed. Expressed milk, donor milk or formula was not explored in the current study and the datasets employed in this study did not contain information regarding donor milk or alternative methods. The survey with women will give a highly optimistic finding as the women who responded to a survey had already a higher rate of intention to breastfeed compared to the routine data population and so the findings from this survey will be biased toward breastfeeding.

In conclusion, intention to breastfeed was the strongest predictor of exclusive breastfeeding outcome at six months. Therefore, more targeted interventions to promote, encourage and maintain breastfeeding duration up to six months need to be implemented early in pregnancy or in pregnancy planning.

## Data Availability

All data produced in the present study are available upon reasonable request to the authors

